# A systematic review of *Schistosoma intercalatum*: an obscure causative agent of human schistosomiasis

**DOI:** 10.1101/2024.08.12.24311882

**Authors:** Oluwaremilekun Grace Ajakaye, Jean Pierre Kambala Mukendi, Eddy Laken Kakiese, Dieudonne Mumba Ngoyi, Masceline Jenipher Mutsaka-Makuvaza

**Affiliations:** Department of Animal and Environmental Biology, Adekunle Ajasin University, Akungba-Akoko, Ondo State, Nigeria; Department of Tropical Medicine, Infectious and Parasitic Diseases, Faculty of Medicine, University of Kinshasa, Kinshasa, Democratic Republic of the Congo; Department of Parasitology, Institut National de Recherche Biomédicale (INRB), Kinshasa, Democratic Republic of the Congo; Department of Microbiology and Parasitology, School of Medicine and Pharmacy, University of Rwanda; National Institute of Health Research, Ministry of Health and Child Care, Zimbabwe

**Keywords:** Schistosomiasis, *S. intercalatum*, hybridization, Biology, Pathology, DRC

## Abstract

**Background:** Human schistosomiasis caused by *Schistosoma intercalatum* is poorly understood compared to other species of public health importance, such as *S. haematobium, S. mansoni*, and *S. japonicum*. The restricted distribution of *S. intercalatum* in Africa, its perceived low virulence, and poor understanding of its pathological consequences are possible reasons for this neglect. However, schistosomiasis as a public health problem cannot be eradicated without adequate knowledge of existing species of schistosomes and the biological interactions between them, their hosts, and the environment. Current information on *S. intercalatum* is often confused with the previous *S. intercalatum* (Lower Guinea strain) which has now been described as *S. guineensis* n. sp. The *S. intercalatum* (Zaire strain) with known foci only in Democratic Republic of the Congo (DRC) is now recognized as the ‘true’ *S. intercalatum* species. Investigators not conversant with the present status of both species still get confused in understanding the literature hence this review. It is essential to discuss available knowledge on *S. intercalatum* and highlight gaps that are critical for deepening our understanding of the parasite in the DRC.

**Methods:** Thus, we systematically review existing available literature on *S. intercalatum* obtained from PubMed Central, Web of Science, Embase, and Scopus databases according to the Preferred Reporting Items for Systematic Reviews and Meta-Analyses (PRISMA) guidelines. Relevant publications were screened and publications reporting *S. intercalatum* from other than the DRC were excluded. Eligible studies on the epidemiology and biology of *S. intercalatum* were reviewed.

**Results:** Based on 38 publications that met our inclusion criteria, we confirm that *S. intercalatum* is the most neglected among *Schistosoma* species responsible for human schistosomiasis. We discuss our synthesis of the available information in the context of the distribution, biology, pathology, and genetics of *S. intercalatum*. Also highlighted are outstanding questions on *S. intercalatum* population genomics, hybridization with other *Schistosoma* species, snail vector host, zoonotic epidemiology, and pathology.

**Conclusion:** The review calls for a rekindling of research interest in the parasite and the need to answer fundamental questions that can deepen our understanding of the parasite epidemiology, disease ecology, and environmental interactions to help in control strategies and elimination.

## Introduction

Three parasitic worm species in the genus *Schistosoma* (*S. mansoni, S. japonicum*, and *S. haematobium*) are the widely recognized and studied causative agents of human schistosomiasis. Schistosoma *japonicum* and *S. mansoni* are responsible for intestinal schistosomiasis, while *S. haematobium* is responsible for urogenital schistosomiasis. Other species include *S. guineensis, S. intercalatum,* and *S. mekongi,* which have a lower global prevalence because of their localized distribution and, thus, are the least studied [1,2].

The adult stages of the parasites lodge in the host’s blood vessels within the intestinal or urogenital systems, where the mature female produces thousands of eggs, which are deposited in various tissues and organs, most notably the liver, bladder, small and large intestine, cervix, and vagina. Most disease pathology arises from inflammatory reactions and immune responses due to trapped eggs in the various organs. It can result in complications such as hepatosplenomegaly, periportal fibrosis, bladder cancer, and female genital schistosomiasis [1, 2].

Although under-studied because of its localized distribution, and first identified in the Upper Congo by Chesterman, *S. intercalatum* is also of medical importance as responsible for rectal schistosomiasis. [3]. In 1934, Fisher [4] described the parasite as a new species of schistosome, and the features of the eggs essentially led to the name *’intercalatum*’ to underscore the intermediate profile when compared to eggs by *S. haematobium* and the bovine schistosome *S. bovis* [4]. Before the year 2003, two unique populations of *S. intercalatum* were recognized and marked by specific geographic locations, snail hosts, prepatent period in snail, egg morphometrics, isoenzyme patterns, Random Amplified Polymorphic DNA (RAPD), and Restriction Fragment Length Polymorphism (RFLP) [5 - 10]. Pages [11] supported by DNA sequencing data, grouped the *S. intercalatum* populations from the Lower Guinea forest region of Cameroon as a unique species described as *S. guineensis* n. sp. Other countries in the Lower Guinea forest region with reported foci for *S. guineensis* include Gabon, Equatorial Guinea, Nigeria, Benin Republic, and Sao Tome and Principe. Presently, *S. intercalatum* is known to have foci only in the Democratic Republic of the Congo (DRC) [12, 13].

Although the distinct taxonomic status of *S. intercalatum* and *S. guineensis* was established in 2003, the large literature on *S. intercalatum* prior to 2003 has yet to be systematically reviewed and sorted according to species. Investigators not conversant with the present status of both species still get confused in understanding the literature. In this review, we focus only on literature pertaining to *S. intercalatum* species with known loci in the DRC. We discuss our synthesis of the available information in the context of the distribution, biology, pathology, and genetics of *S. intercalatum*.

## Methodology

### Eligibility

Our primary objective was to compile peer-reviewed research publications, review articles, and case reports pertaining only to *S. intercalatum* species from the DRC. We aimed at synthesizing the information from all available publications from the 1900s till date. Our inclusion and exclusion criteria were based on the geographical source of the *S. intercalatum* reported in a publication. Literature reporting *S. intercalatum* originating from the DRC was included, while publications citing *S. intercalatum* originating from other African countries were considered as *S. guineensis* and thus excluded from our pool of literature for the review.

### Literature search strategy

Three authors, OGA, JPKM, and ELK were responsible for the literature search and selection. Both JPKM and ELK are fluent French speakers and were therefore able to thoroughly appraise publications that were in the French language. A literature search of bibliographic databases Web of Science, EMBASE, SCOPUS, and PubMed Central was conducted between April 2022 and August 2023 for publications reported between the year 1900 to August 2023. To complement the database search, a manual search of publications was conducted by examining references from reviews and opinion articles on *S. intercalatum* or other *Schistosoma* species. Two search terms, “*Schistosoma intercalatum*” and “*intercalatum*” were used within field tags ‘Article Title,’ Abstract,’ and ‘Key Words’. Analysis of the retrieved literature complied with the reporting elements for systematic reviews guidelines prescribed in the PRISMA 2020 statement for reporting systematic reviews [14].

## Results

A total of 2,814 publications were retrieved from bibliographic databases, and after screening, 38 met the eligibility criteria and were used for full-text review (Fig.1).

**Fig.1.**
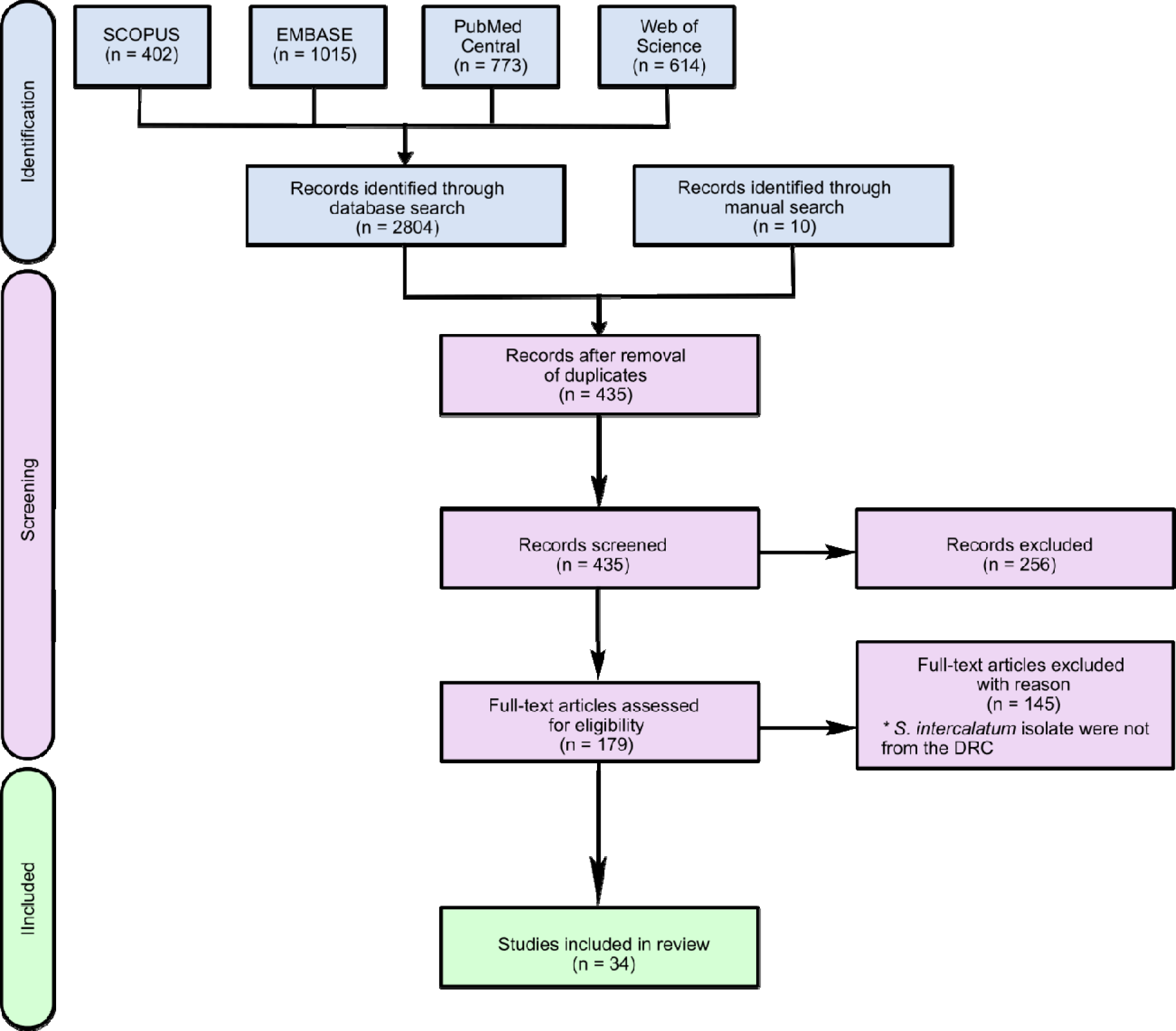
PRISMA chart of literature selection process on *Schistosoma intercalatum*

## Discussion

### Schistosoma intercalatum in the DRC

The history of *S. intercalatum* in the DRC dates to early 1900 in the regions around the former Belgian Congo (the present-day countries of DRC, the Republic of Congo, and parts of the Central African Republic). Several investigators reported terminal-spined *S*. *haematobium*-like eggs exclusively from stool samples in several foci in the Belgian Congo (see review in Fisher, [4]). Gillet and Wolfs [15] have also provided detailed historical accounts of the distribution and biology of three agents of schistosomiasis (*S. mansoni*, *S. haematobium*, and *S. intercalatum*) in the Belgian Congo. The last review of published research on schistosomiasis in the DRC confirmed the occurrence of one or more species of *Schistosoma* by microscopy (*S. mansoni*, *S. haematobium*, and *S. intercalatum*) in all but one of the provinces in the country [16]. The current prevalence rate of *S. intercalatum* is unknown. However, old reports based on microscopic analysis indicated it ranges between 1 and 10% [9, 17], except for the study by Gillet and Wolfs [15] and Schwetz [18], which reported above 30%. Although there are no recent reports of the bovine schistosome *S. bovis* in the DRC, its occurrence has been documented previously [18, 19]. In these studies, which used 781 cattle to determine the possible associations between different bovine trematode species in the Ituri region of the DRC, *S. bovis* adult worms were recovered from the mesenteric veins.

### Biology of S. intercalatum

#### i. Lifecycle

*Schistosoma intercalatum* lifecycle, larval and adult stages are similar to other known human *Schistosoma* species (Fig. 2). Fisher [4] and Wright [5] have provided morphometric and behavioral information on the species from isolates originally derived from a patient in Kinshasa, which were experimentally passaged through infected mice or hamsters.

**Figure 2.**
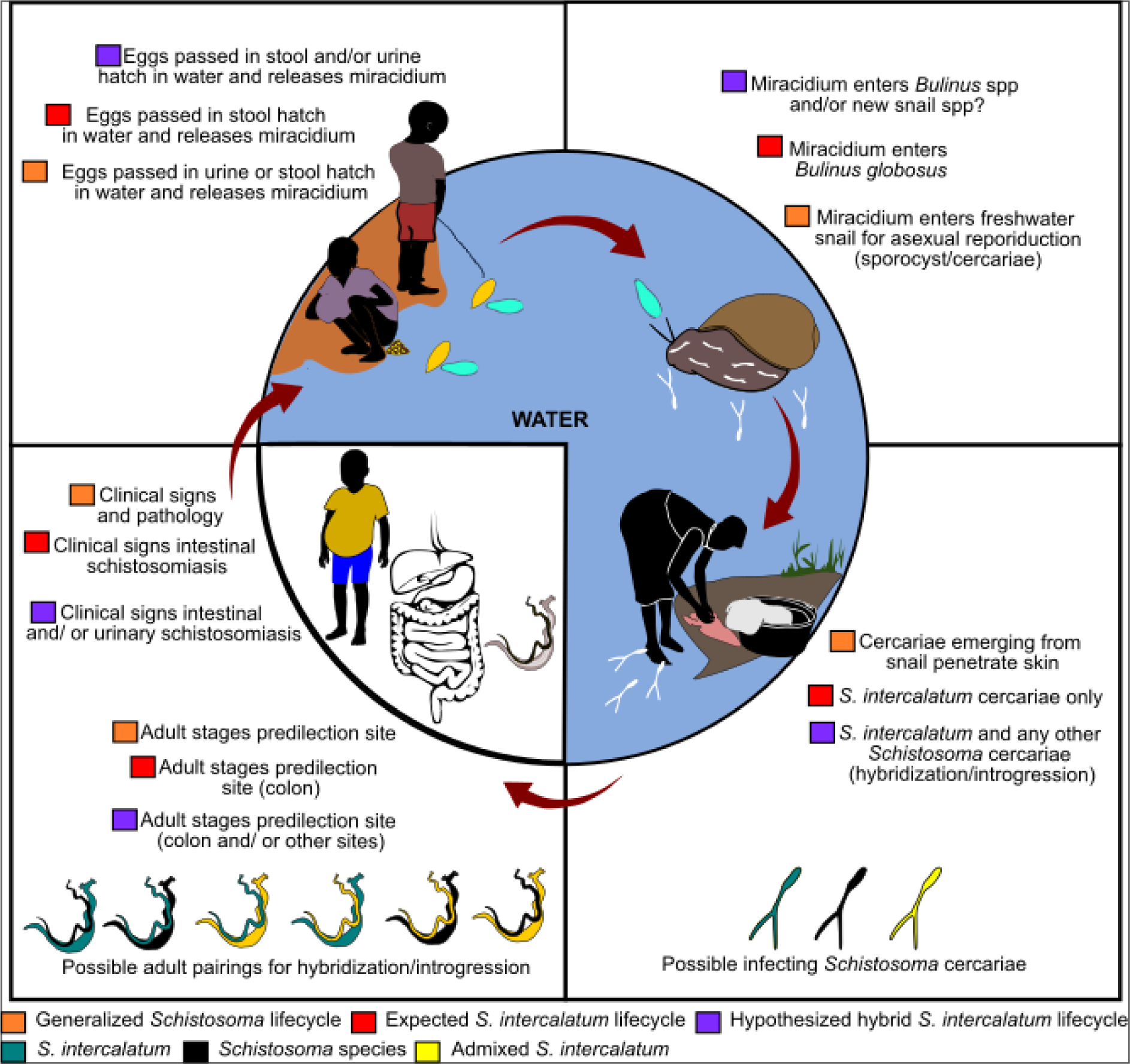
The life-cycle of *S. intercalatum* and its hybrid.

The cercariae of *S. intercalatum* and *S. guineensis* (formerly *S. intercalatum*, Lower Guinea strain) have been observed to concentrate near the surface of the water and readily attach to floating debris or the sides of a container in the laboratory, in contrast to *S. mansoni* cercariae, which typically swim to the surface of the water, rest briefly, and sink partially before swimming back [5, 20].

#### ii. Snail host

Crucial to the lifecycle of schistosomes are the appropriate intermediate snail hosts, and for *S. intercalatum*, Fisher [4] reported *Physopsis africana* (old nomenclature), now *Bulinus africanus*. However, the known difficulty in accurately morphologically identifying *Bulinus* species in Africa warrants the use of currently available molecular tools to aid species identification [8, 21 - 23]. Wright et al. [5] were unsuccessful in infecting several *Bulinus* species (*B. africanus*, *B. globosus*, *B. ugandae*, *B. nasutus*, *B. forskalii*, and *B. camerunensis*) that had been collected from various countries in Africa and bred in the lab with *S. intercalatum*. Only *B. globosus* from South Africa, Ghana, Zimbabwe, Sudan, and *B. wrighti* from Saudi Arabia were successfully infected. However, Wright et al. [5] observed a lengthier prepatent period compared with other species of *Schistosoma* investigated in the study. Frandsen [6, 24], however, demonstrated successful experimental *S. intercalatum* infections of *B. globosus* from Malawi, Togo, Rhodesia (Zimbabwe), and DRC. Interestingly, *B. africanus* from Kenya and Tanzania were susceptible to *S. intercalatum*, while the albino strain of *B. globosus* from Zimbabwe was the most compatible compared to other snail species successfully infected. De Clercq [17] also confirmed *B. globosus* as natural hosts for *S. intercalatum* in the DRC. However, Tchuem Tchuenté et al. [9] were unsuccessful in finding naturally infected snails for *S. intercalatum* in the DRC but reported similar experimental results reported by Wright et al. [5]. The snail infection trials demonstrated incompatibility between *S. intercalatum* and *B. forskalii*, *B. truncatus/B. tropicus* complex, and *B. africanus* group, except for *B. globosus* from Zambia and *B. wrighti* snails. Although Frandsen [6, 24] has done extensive snail host specificity experiments for *S. intercalatum*, they were by no means exhaustive of the diversity of *Bulinus* species in Africa [8]. Also, the susceptibility and compatibility of other non-*Bulinus* species to *S. intercalatum* are yet unknown.

How snail hosts genetic diversity impacts *S. intercalatum* presents an intriguing line of research, and in the scenario of hybridization between *S. intercalatum* and other *Schistosoma* species, it would be essential to determine if hybrid *S. intercalatum* recruits new snail hosts. Questions on the genetic basis of *Bulinus*-*S. intercalatum* interaction is unanswered. On the possibility of control strategies targeting snail hosts, there is a need to identify genes that convey natural resistance to *S. intercalatum* in *Bulinus* species, which may facilitate ways of disrupting the parasite’s life cycle and reducing transmission to humans.

#### iii. Reservoir hosts

Unlike the two main agents of intestinal schistosomiasis (*S. mansoni* and *S. japonicum*) with a broad spectrum of reservoir hosts, no known natural reservoir hosts for *S. intercalatum* in the DRC have been identified. Monkeys, Congo clawless otters (*Aonyx congica*), sheep, and goats examined for *S. intercalatum* by Fisher [4] were uninfected. However, Fisher [4] successfully infected a male sheep with cercariae of *S. intercalatum*. Schwetz [18] was also able to infect both sheep and goats with *S. intercalatum* using the same route of infection as by Fisher [4], although no one else has ever tried this route to confirm these findings.

The role of animal reservoir hosts in *S. intercalatum* transmission remains unexplored. Fisher [4] and Schwetz [18] examined a few mammalian species without recording any natural host for *S. intercalatum.* Several species of rodents across Africa are known to serve as natural reservoir hosts of several species of *Schistosoma* and hybrid *S. haematobium* x *S. bovis* [25, 26]. The rodent species frequenting water bodies in schistosomiasis endemic communities in the DRC will be interesting for investigation. There is now a wide range of molecular approaches for detecting pathogens that do not require invasive sampling. Also, environmental DNA techniques provide an opportunity for detecting nucleic acid traces in various environmental samples [27 - 29].

### Pathology of S. intercalatum

There is generally a dearth of information on the clinical aspects of *S. intercalatum* due to limited studies on the parasite. There is a need for an adequate understanding of the pathological impacts of the parasite to inform necessary control and elimination strategies. Fisher [4] and Schwetz [18] have highlighted several *S. intercalatum* clinical signs similar to other intestinal human schistosomiasis caused by *S. mansoni*, S*. guineensis*, and *S. japonicum*. Wolfe [30] provided a case report of an American family returning to the USA from Kisangani, Eastern DRC who were infected with *S. intercalatum* after parasitological examination of stool. The common symptoms that have been reported include abdominal pain, blood in stool, dysentery, and a few cases of palpable, enlarged livers. None of the reported cases showed any marked degree of anemia.

### Molecular epidemiology of *S. intercalatum*

Before the separation of the Lower Guinea strain of *S. intercalatum* (now *S. guineensis*) from the DRC *S. intercalatum* species, genotyping studies of both species were performed using isoenzyme patterns, Random Amplified Polymorphic DNA (RAPD) and Restriction Fragment Length Polymorphism (RFLP) [7 - 9]. DNA-sequence data based on three mitochondrial genes, (cytochrome oxidase subunit 1 (cox1), NADH dehydrogenase subunit 6 (nad6) and small ribosomal RNA (rrnS)) was pivotal evidence for the eventual distinctions between the two species and the subsequent description of *S. guineensis* [10 - 12]. Presently the available DNA sequence data on *S. intercalatum* archived on the NCBI nucleotide database is problematic because some of the sequences labeled as *S. intercalatum* are that of *S. guineensis* [11 - 13] (Table 1). Unfortunately, most nucleotide sequences deposited at the NCBI are rarely revised or corrected after their initial submission. Apart from sequence alignments consistent with the phylogram reported in [11, [12], and [13], users can quickly ascertain the NCBI sequence data on *S. intercalatum* by checking the country of origin of the isolate (*S. intercalatum* will expectedly be from DRC). Webster et al. [13] have also provided information for the correct taxonomic names for sequences on the NCBI nucleotide database labeled as *S. intercalatum* (now *S. guineensis*) and those from DRC representing *S. intercalatum*.

**Table 1.** Correct names for nucleotide sequences labeled as *Schistosoma intercalatum* on NCBI.

| Source country | Name on NCBI | Accession # (isolate) | Gene | Base pairs | Reference | Correct name |
| --- | --- | --- | --- | --- | --- | --- |
| DRC | *S. intercalatum* | DQ354362,<br>DQ354363,<br>DQ354364<br>(NHM3367/<br>NHM3397)<sup>+</sup> | 28S<br>rRNA, 18S<br>rRNA,<br>cox1, | 3834,<br>1855,<br>1126 | [13] | *S. intercalatum* |
| São Tomé and Príncipe | *S. intercalatum* | AY157208,<br>AY157235,<br>AY157262<br>(NHM3188) | cox1, 18S<br>rRNA, 28S<br>rRNA | 1125,<br>1863,<br>3834 | [31] | *S. guineensis* |
| DRC | *S. intercalatum* | JX844184 &<br>JX844185 | tetraspanin<br>-23 (TSP-23) | 1580 | [32] | *S. intercalatum* |
| Cameroon | *S. intercalatum* | U22160,<br>U22166 | cox1, 5.8S,<br>ITS2, 28S<br>rRNA | 372,<br>331 | [33] | *S. guineensis* |
| DRC | *S. intercalatum* | DQ831703,<br>DQ831704** | clone<br>Sinter23 &<br>Sinter25<br>repetitive<br>sequences | 493 &<br>503 | [34] | *S. intercalatum*<br>& *S. guineensis* |
| DRC | *S. intercalatum* | AJ519529,<br>AJ519515,<br>AJ416894,<br>AJ419779<br>(held in<br>France) | 28S rRNA,<br>cox1, nad6,<br>12S rRNA | 680,<br>1224,<br>420,<br>754 | [12] | *S. intercalatum* |
| DRC | *S. intercalatum* | AJ519527,<br>AJ519519,<br>AJ416899,<br>AJ419781<br>(NHM3396) | 28S rRNA,<br>cox1, nad6,<br>12S rRNA | 680,<br>1224,<br>420,<br>754 | [12] | *S. intercalatum* |
| São Tomé<br>and<br>Príncipe** | *S. intercalatum* | Z72146 | 18S rRNA | 333 | Unpublished | *S. guineensis* |
| Cameroon | *S. intercalatum* | U42564,<br>U42558 | 18S rRNA<br>v4 domain,<br>28S rRNA<br>D1 domain | 314,<br>173 | [35] | *S. guineensis* |
| Cameroon | *S. intercalatum* | L03655 | 16S rRNA | 286 | [36] | *S. guineensis* |
| DRC | S.<br>*intercalatum* | GCA_94447<br>0365.2*** | Reference<br>genome | 386,<br>052,<br>197 | Unpublished** | S.<br>*intercalatum* |

DRC (Democratic Republic of the Congo), NHM (Natural History Museum, London), Conflicting isolate labels in publication and NCBI\*. Unclear which geographical location is associated with each sequence\*\*, Based on Webster et al., 2006\*\*, Current version of submitted GenBank assembly\*\*, Originally sequenced as part of International Helminth Genomes Consortium [31].

Table 1. Correct names for nucleotide sequences labeled as *Schistosoma intercalatum* on NCBI

### Outstanding questions on *S. intercalatum*

Although schistosomiasis is considered a neglected tropical disease, *S. intercalatum* appears to be the most neglected among the species that cause human schistosomiasis. It is probably due to the difficulty in accessing new specimens from DRC after decades of protracted conflicts and insecurity in the country. The genetic basis for the restricted distribution of *S. intercalatum* is still unknown. However, the availability of modern DNA sequencing technologies, including Next Generation Sequencing (NGS), provides an opportunity for robust genomic investigations on *S. intercalatum*. Some outstanding questions still in need of answers include:

i. **Genetic diversity and population genetics / genomics**: Compared to other *Schistosoma* species of medical importance, there is no information on the genetic diversity or population genetics of *S. intercalatum*. Given the large landmass area of the DRC with its diverse ecological zones and abundant wildlife, it would be interesting to unravel the evolutionary dynamics of the parasite over time.
ii. **Hybridization**: Interspecific hybridization has been demonstrated to occur in the laboratory within the *S. haematobium* species complex, which is comprised of eight sister species including *S. intercalatum*, *S. guineensis*, *S. bovis*, *S. mattheei*, *S. margrebowei*, *S. leiperi*, *S. curassoni,* and *S. kisumuensis)* [37, 38]. Increasingly, hybrids are being recovered from field sites, but the true extent to which this is occurring in nature remains enigmatic. This is largely because no systematic genotyping strategy exists, and only limited gene markers or genome-scale datasets have been derived from *Schistosoma* samples collected from natural infections. To date, interspecific hybrids within the *S. haematobium* group species have been described especially between *S. haematobium x S. bovis* (reviewed in [39]), *S. haematobium x S. guineensis* [38], *S. haematobium x S. mattheei* [39] and *S. bovis x S. curassoni* [40]. DNA sequence data has thus far not identified any field isolates that exist as *S. intercalatum* hybrids with other *Schistosoma* species, although, based on isoenzyme analysis, Tchuem Tchuenté et al. [9] reported that natural hybridization is occurring between *S. haematobium* and *S. intercalatum* in the DRC. The reference genome for *S. intercalatum,* sequenced as part of a consortia project [41] is freely available on the NCBI Short Read Archives (see [40] for details) and appears to be devoid of mixed ancestry. However, previous *S. intercalatum* hybridization experiments with *S. guineensis* or *S. mansoni* have established that *S. intercalatum* is mating competent under laboratory conditions with its sister species, *S. guineensis* [42 - 44]. Through a suite of genome analysis tools, it is now possible to analyse the direction of introgression within inter species hybrids, to identify introgressed genes, and to establish whether the hybrids could alter virulence phenotypes [37, 45].
iii. **Epidemiology and pathology**: Tchuem Tchuenté [9], to our knowledge, is the last peer-reviewed publication on the epidemiology of *S. intercalatum* from the DRC. There is a need for a nationwide epidemiological survey to determine the current prevalence of *S. intercalatum* or its hybrid in the DRC and neighbouring countries and detailed documentation of clinical signs of the disease. Importantly, molecular studies of samples from such nationwide surveys will provide information to understand if *S. intercalatum* has been entirely replaced by *S. haematobium* as was reported for *S. guineensis* replacement by *S. haematobium* in [46 - 48].

## Conclusion

Arguably, the 1990s intense security challenges in the DRC affected the study of *S. intercalatum*. Now that the security in the DRC has improved, there is a need for renewed interest in studying *S. intercalatum*. While lessons learned from current research in Africa on the two major African human schistosome species, *S. mansoni* and *S. haematobium*, can be applied in other endemic regions, there is no such scenario for *S. intercalatum*. It is of epidemiological importance to know the degree to which *S. intercalatum* is cross-hybridizing vs. inbreeding and whether this is impacting the incidence and severity of rectal schistosomiasis in the country. In light of the World Health Organization’s roadmap for the elimination of human schistosomiasis, an understanding of the molecular epidemiology of all causative agents and the complex interactions between them, their various host species, and the disease ecology are all vital.

## Declarations

### Ethics approval and consent to participate

Not Applicable

### Consent for publication

Not Applicable

### Availability of data and materials

All data generated or analysed during this study are included in this published article.

### Author Contributions

OGA, ELK and JPKM did the literature searches; OGA and JPKM wrote the manuscript. JPKM and MJM updated the text; ELK and DMN reviewed the manuscript; and OGA and JPKM organized and edited the final manuscript.

## Data Availability

All data produced in the present work are contained in the manuscript

## Acknowledgements

We would like to thank Elisha Enabulele for generating the outstanding figure, and Michael Grigg for critical reading of the manuscript.

